# Comparison of Machine Leaning Models for Prediction of Acute Pain Severity and On-Treatment Opioid Utilization in Oral Cavity and Oropharyngeal Cancer Patients Receiving Radiation Therapy: Exploratory Analysis from a Large-Scale Retrospective Cohort

**DOI:** 10.1101/2024.02.06.24302341

**Authors:** Vivian Salama, Laia Humbert-Vidan, Brandon Godinich, Kareem A. Wahid, Dina M. ElHabashy, Mohamed A. Naser, Renjie He, Abdallah S.R. Mohamed, Ariana J. Sahli, Katherine A. Hutcheson, Gary Brandon Gunn, David I. Rosenthal, Clifton D. Fuller, Amy C. Moreno

## Abstract

**Background:** Acute pain is a common and debilitating symptom experienced by oral cavity and oropharyngeal cancer (OC/OPC) patients undergoing radiation therapy (RT). Uncontrolled pain can result in opioid overuse and increased risks of long-term opioid dependence. The specific aim of this exploratory analysis was the prediction of severe acute pain and opioid use in the acute on-treatment setting, to develop risk-stratification models for pragmatic clinical trials.

**Materials and Methods:** A retrospective study was conducted on 900 OC/OPC patients treated with RT during 2017 to 2023. Clinical data including demographics, tumor data, pain scores and medication data were extracted from patient records. On-treatment pain intensity scores were assessed using a numeric rating scale (0-none, 10-worst) and total opioid doses were calculated using morphine equivalent daily dose (MEDD) conversion factors. Analgesics efficacy was assessed based on the combined pain intensity and the total required MEDD. ML models, including Logistic Regression (LR), Support Vector Machine (SVM), Random Forest (RF), and Gradient Boosting Model (GBM) were developed and validated using ten-fold cross-validation. Performance of models were evaluated using discrimination and calibration metrics. Feature importance was investigated using bootstrap and permutation techniques.

**Results:** For predicting acute pain intensity, the GBM demonstrated superior area under the receiver operating curve (AUC) (0.71), recall (0.39), and F1 score (0.48). For predicting the total MEDD, LR outperformed other models in the AUC (0.67). For predicting the analgesics efficacy, SVM achieved the highest specificity (0.97), and best calibration (ECE of 0.06), while RF and GBM achieved the same highest AUC, 0.68. RF model emerged as the best calibrated model with ECE of 0.02 for pain intensity prediction and 0.05 for MEDD prediction. Baseline pain scores and vital signs demonstrated the most contributed features for the different predictive models.

**Conclusion:** These ML models are promising in predicting end-of-treatment acute pain and opioid requirements and analgesics efficacy in OC/OPC patients undergoing RT. Baseline pain score, vital sign changes were identified as crucial predictors. Implementation of these models in clinical practice could facilitate early risk stratification and personalized pain management. Prospective multicentric studies and external validation are essential for further refinement and generalizability.

## Introduction

Acute pain is one of the most common debilitating symptoms that develops during Radiation Therapy (RT), in oral cavity and oropharyngeal (OC/OPC) cancers. For OC/OPC, the standard RT is given in multiple fractions per day, for 5-7 weeks of RT alone or with concurrent chemotherapy [1–4]. Despite the improved OC/OPC patients’ outcome with the implementation of advanced adaptive RT techniques, several acute adverse symptoms are often reported during and after RT [5]. These adverse symptoms have a negative impact on the patients’ quality of life (QoL) [6]. Acute mouth/throat pain has been reported to affect over 90% of OC/OPC patients, with up to 80% requiring an opioid prescription to manage cancer and/or treatment-associated pain [7, 8].

Despite the availability of the World Health Organization’s (WHO) “analgesic ladder” guideline designed for pain relief [9–11], managing pain, particularly in patients undergoing RT for head and neck cancers (HNC), remains a significant challenge for healthcare providers, with nearly one-third of HNC patients continuing to experience severe, uncontrolled pain [12–14]. The challenge in the control of RT-induced acute pain is potentially related to the complex nature of pain, the multifactorial etiology of pain and the different response rates of individuals to pain treatment [5, 15]. Most healthcare professionals prescribe opioids for pain control during RT in OC/OPCs [8, 12, 16]. The escalated doses of opioids for acute pain management during RT contribute to heightened rates of morbidity and raises concerns about potential opioids’ side effects and substance abuse [17]. These challenges in pain management not only complicate care but also have a detrimental impact on the QoL for the survivors within this cancer population. Approximately 45% of long-term HNC survivors report chronic pain, with more than 10% exhibiting severe chronic pain with chronic opioid usage [18]. The long-term opioid usage raises the risks of opioid dependence and drug addiction which may lead to patient death [19–21]. The overuse of high doses of opioids to address acute pain during RT adds to the complexity of the patients’ care and poses risks that may exacerbate their overall health outcomes and well-being [22].

Artificial Intelligence and machine learning (AI/ML) models are currently being explored for their potential use in risk stratifying patients according to various risks, especially in the domain of pain medicine and opioid usage [23]. These models aim to optimize pain management and assist in personalized treatments through risk stratification and decision-making [23]. For example, Chao et al, used ML algorithms to identify chest wall pain induced by RT in non-small cell lung cancer (NSCLC) patients treated with Stereotactic Body Radiation Therapy (SBRT) [24] and Olling et al., generated ML predictive models for predicting pain while swallowing (odynophagia) during RT in NSCLC [25]. While approximately 44 studies exploring ML models to predict cancer pain, no studies investigated the role of ML models in pain prediction in head and neck cancers (HNC) and how they can aid in guiding decisions related to the use of opioids in these individuals.

The primary objective of the ongoing study is to address this gap in knowledge by a) comparing the performance of various ML algorithms as predictive models for predicting acute pain levels, b) projecting opioid doses at the end of RT in OC/OPC patients and c) identifying the importance of relevant clinical predictors in classifying acute pain and predicting the required opioid dosages.

## Materials and Methods

### Patient data

A retrospective study was conducted using a cohort of oral cavity, oropharyngeal cancer and unknown primary cancer patients treated with RT at our institution from 2017 to 2023. The study has been approved by The University of Texas MD Anderson Cancer Center (MDACC) Institutional Review Board (IRB) (2024-0002). Since most unknown primary cancers end up being oropharyngeal cancer (OPC) or oral cavity cancer (OC), they were included in our study.

Eligible OC/OPC patients for inclusion included those with a pathologic diagnosis of squamous cell carcinoma (SCC) treated with RT or chemoradiation therapy (CRT). RT modalities included photons (e.g., intensity-modulated radiotherapy (IMRT), volumetric modulated arc therapy (VMAT)) and proton therapy (i.e., IMPT).

#### Predictors

Clinical data extracted from the electronic health record system included patient demographics, exposure/social history (smoking, alcohol, drug abuse), tumor and staging characteristics, cancer therapy details (systemic therapy, surgery, RT), vital signs (weight and heart rate), medications, and baseline and last on-treatment visit [i.e., weekly see visit (VSV)] acute pain scores. Delta changes in weight were calculated using the equation: [(last WSV weight-baseline weight)/baseline weight] *100. Categorical encoding was applied to the relevant variables and robust scaler normalization of normally distributed continuous variables was implemented.

#### Outcome

Desired output variables for the ML models included end-of-RT pain scores and opioid usage, the latter defined as the total morphine equivalent daily dose (MEDD). Pain intensity was documented during nursing visits using a numerical scale from 0 (none) to 10 (as bad as you can imagine). Our output pain intensity variable was dichotomized into two classes, non-severe pain (0-6) and severe pain (7-10). This cutoff was used based on available literature [5, 26, 27] and current physician considerations of a pain score of 7 or greater representing high risks for uncontrolled pain that need readdressing of applied pain control regimens [28, 29]. The other endpoint (MEDD) at the last WSV was calculated using the Centers for Disease Control and Prevention (CDC) guidelines [30–32]. The total MEDD was calculated as follows: the unit dose of all opioids (i.e., tramadol, hydrocodone, oxycodone, morphine, methadone, transdermal fentanyl) prescribed during the last WSV were collected and multiplied by the prescribed frequency (i.e., twice per day) and their CDC-based MEDD conversion factors (hydrocodone= 1, hydromorphone= 4, morphine= 1, oxycodone= 1.5, tramadol= 0.1, transdermal fentanyl= 2.4 and methadone according to the dose (1-20 mg/day=4, 21-40 mg/day=8, 41-60 mg/day= 10 and >=61-80 mg/day =12)). The total MEDD was then calculated by summing all opioid specific MEDDs. As a desired output variable, total MEDD was dichotomized into two classes, low MEDD (total MEDD <50 mg/day) and high MEDD (total MEDD >=50 mg/day). We selected 50 mg/day as a cut off given the mean total MEDD of our cohort (52mg) and based on CDC guidelines [23–25]. The third endpoint considered was the status of analgesic efficacy based on the pain intensity and the total MEDD at the end of RT. Analgesic efficacy variable was dichotomized into two classes: analgesic efficacy (non-severe pain <7 and low total MEDD <50) and analgesic inefficacy (severe pain >=7 and high total MEDD >=50). Severe pain and high doses of opioids showed increased disability risks, poor outcomes and low QOL, thus it is essential to identify analgesic efficacy in head and neck cancer patients pre-emptively for better management of these patients during RT [19, 20, 29].

### Descriptive Statistics

Differences in patient characteristics between pain classes and total MEDD were compared using the Chi-square test (the Likelihood Ratio) for categorical variables and Wilcoxon test for numeric variables. A 2-sided P-value less than 0.05 was considered statistically significant.

### Classification models

The full dataset was randomly split after stratification, into training dataset (70%) and testing dataset (30%). Pre-processing of training dataset was conducted separately from the testing dataset. Pre-processing included: 1) categorical variables elements were converted into numerical values. 2) Patients with any missing value were dropped from the dataset, the number of missing data was calculated for each variable (if a variable has > 10% missing data, the variable would be excluded). 3) Normalization of numeric variables using robust scaler if normal distribution of the variable and there are outliers [33].

#### Model training

Fifteen features were used to build the models as input variables. Four ML classification models were trained and included Logistic Regression (LR), Support Vector Machine (SVM), Random Forest (RF), and Gradient Boosting Models (GBM). Hyper-parameter optimization was performed with manual grid search. The LR model was initialized with default settings, using scikit-learn function of Logistic Regression: sklearn.linear_model.LogisticRegression. (penalty=’l2’, C=1.0, solver=’lbfgs’, max_iter=100, fit_intercept=True, random_state=None). The RF model, comprised of 100 decision trees, the function used was: sklearn.ensemble.RandomForestClassifier (n_estimators=100, random_state=10). For predicting MEDD, this python function was used RandomForestClassifier(n_estimators=100, random_state=10, max_depth=3, min_samples_leaf=3). For building the GBM, we used scikit-learn function: sklearn.ensemble.GradientBoostingClassifier (n_estimators=100, learning_rate=0.1, max_depth=2, random_state=12) and for predicting total MEDD, we used scikit-learn function: GradientBoostingClassifier (n_estimators=100, learning_rate=0.1, max_depth=2, random_state=12, min_samples_split=3).

#### Model evaluation

The dataset was split into a training (70%) and a test dataset (30%) with stratified sampling using a ten-fold cross-validation (CV) approach. Model performance was assessed on the test dataset in terms of discrimination performance and model calibration. The discriminative ability was measured using the following metrics: area under the receiver operating curve (AUC), recall, precision, and F1 score. Differences in AUC scores between models were assessed using the DeLong test, R pROc package was used for the DeLong test [34, 35]. Calibration performance was assessed with the calibration curve, which visually depicts the relationship between the predicted probabilities of the positive class and the observed fraction of positive instances. This curve allows for a qualitative assessment of how well the predicted probabilities align with the true outcomes. Additionally, the Expected Calibration Error (ECE) was computed for each model [36, 37]. ECE quantifies the average discrepancy between the predicted probabilities and the observed frequencies, providing a scalar measure of calibration performance. Specifically, ECE was calculated as the mean absolute difference between the observed and predicted probabilities across predefined bins. A lower ECE value indicates better calibration [36].

#### Feature importance

The determination of feature importance for the highest performing model (i.e., the highest AUC) was computed differently according to the model, to elucidate the individual contributions of each predictor variable to the model’s overall performance. For the GBM and RF models, feature importance was calculated through bootstrapped resampling and the calculation of both mean and standard deviation across 100 runs. The mean importance reflected the average contribution of each feature, while the standard deviation (SD) provided insights into the variability and uncertainty associated with these contributions. For the LR and the SVM classifiers, evaluation of feature importance was conducted through the application of permutation feature importance analysis. The resulting feature importance values were then sorted and visualized using a boxplot, providing a comprehensive view of the distribution of feature importance.

Scikit-learn packages were used for ML modeling, validation, and evaluation. All statistical analyses were performed by python 3.12, JMP PRO 15 and R studio version 4.0.5.

## Results

### Patients Characteristics

A total of 900 patients with OC (n=100, 11%), OPC (n=772, 86%) or unknown primary (n=28, 3%) were included in our study Characteristics. Table 1 provides a summary of the cohort characteristics and the results of the Chi-square and Wilcoxon tests.

**Table 1:**
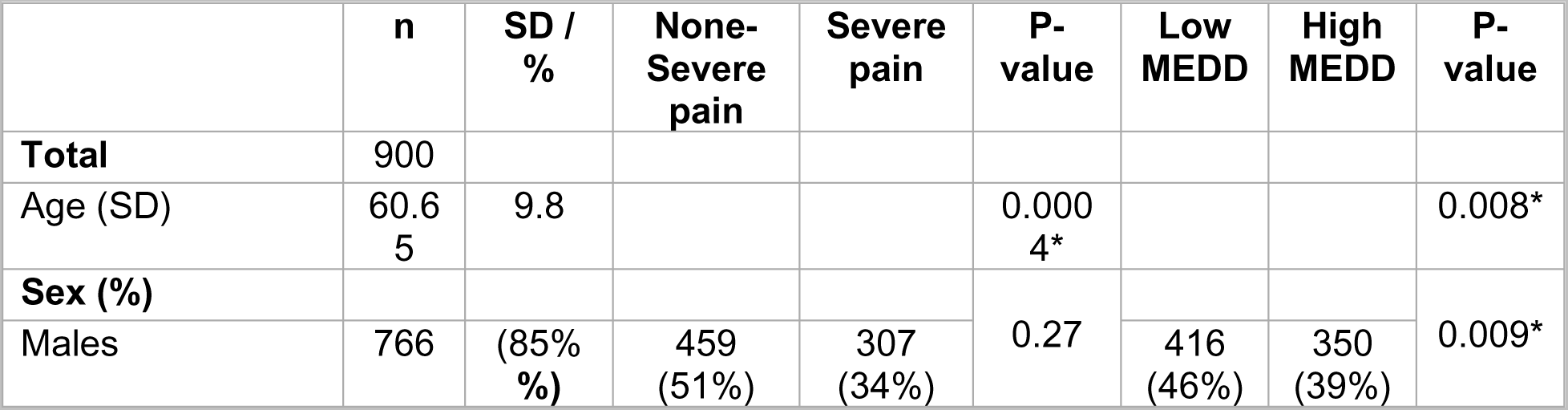
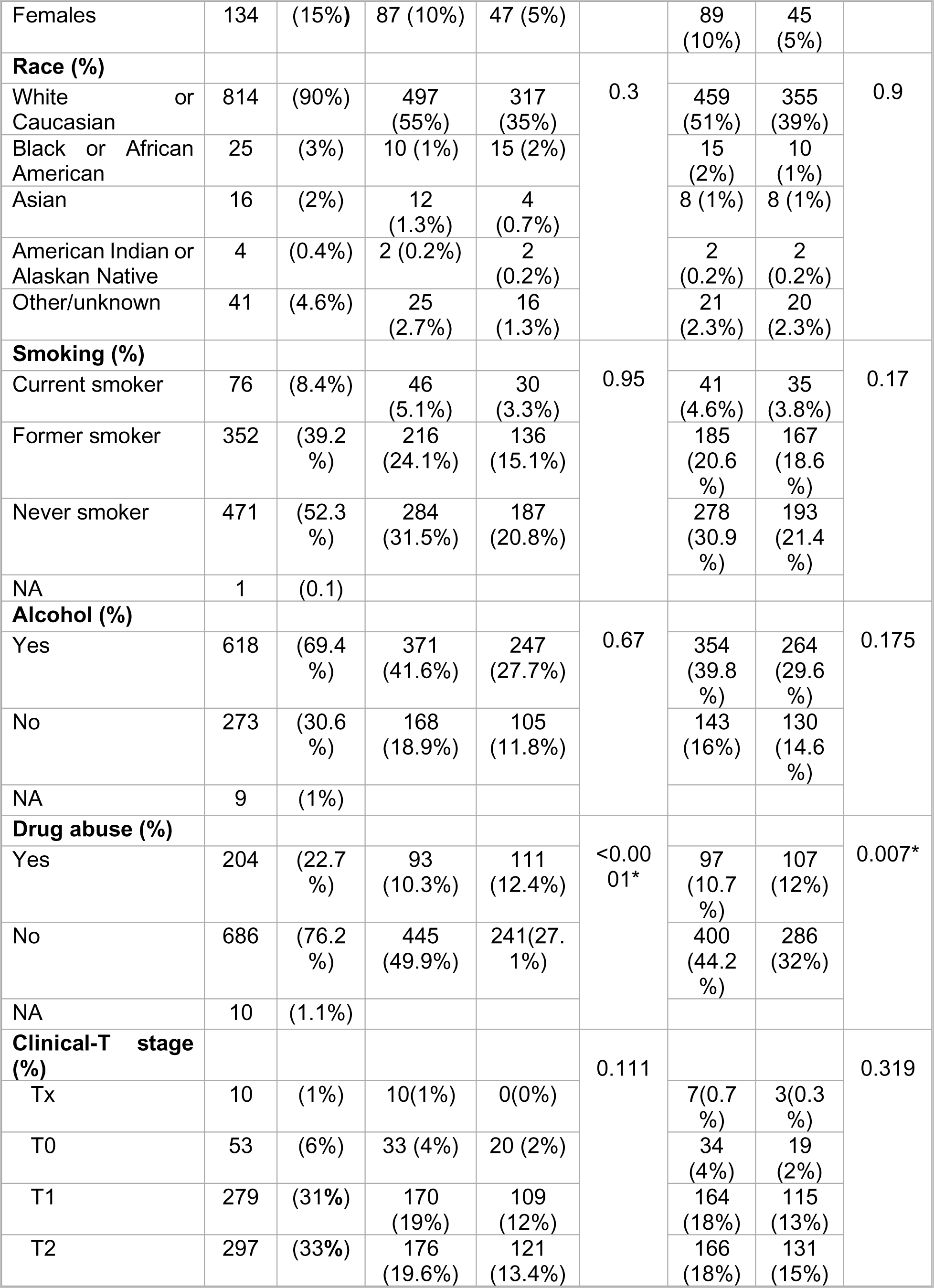
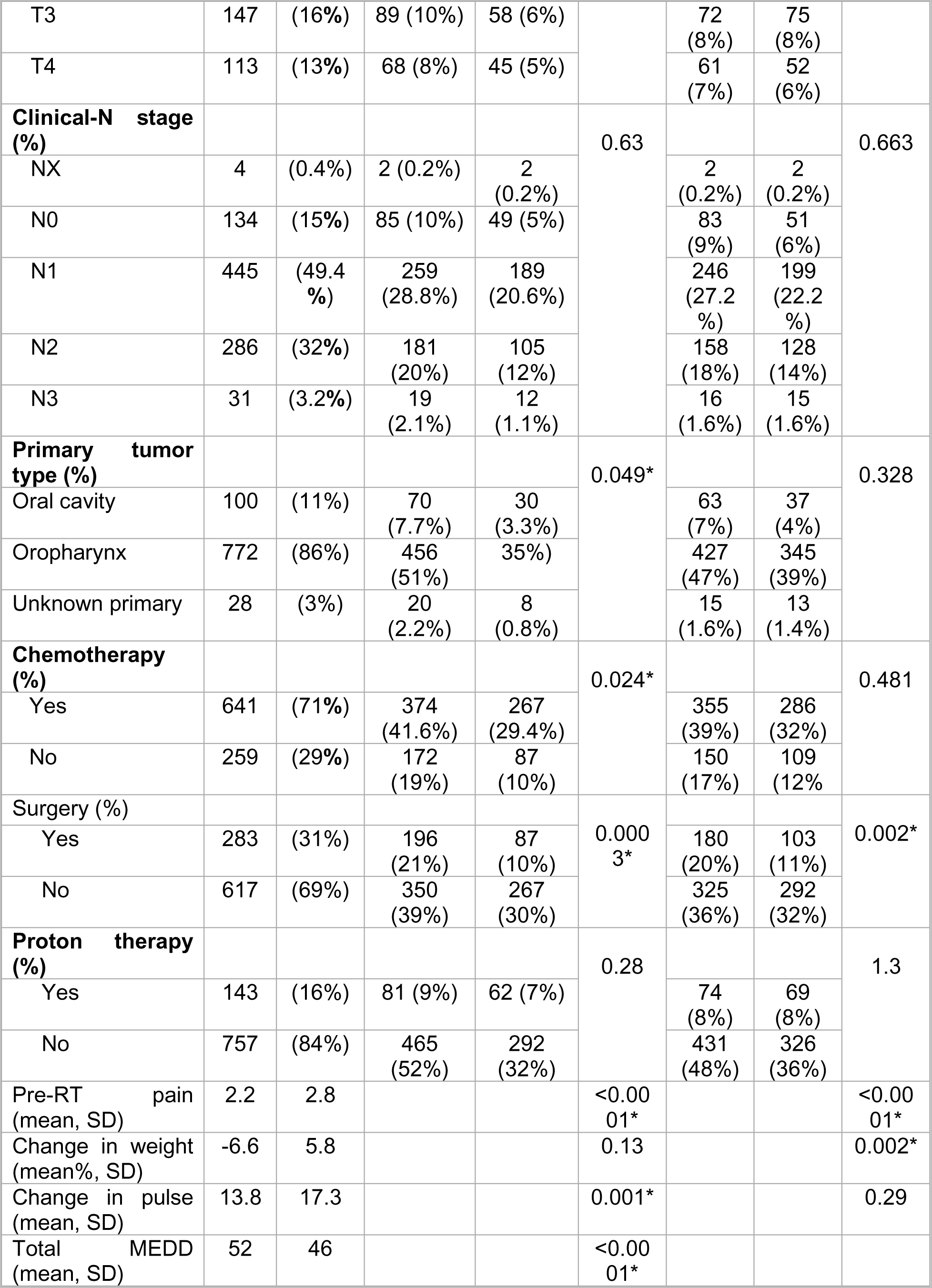
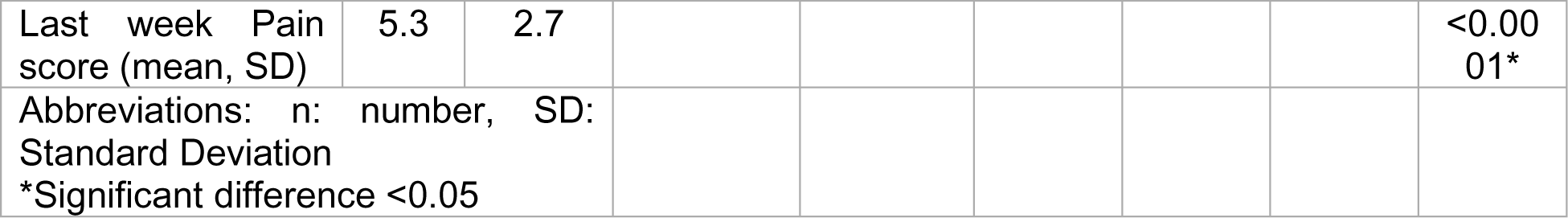
Patients characteristics stratified by acute pain intensity and total MEDD.

### Models’ development and comparison for predicting acute pain intensity at the end of RT in OC/OPC

A total of 838 patients, non-severe pain (n= 502, 60%), severe pain (n=336, 40%), were included in this analysis after dropping patients with missing data. Results of discrimination metrics were summarized in Table 2. The results of the four ML models for predicting pain intensity by the end of RT showed that Gradient Boosting model had the highest, AUC (0.71) Recall (0.39) and F1 score (0.48), indicating that GB, with its well-rounded performance across multiple metrics, appears particularly promising for risk stratification and prediction of acute pain intensity by the end of RT in OC/OPC. The AUC of RF, SVM and LR models were 0.69, 0.65 and 0.64 respectively. However, no significant difference was detected in AUC scores between different models (Figure 1.a). DeLong test results were summarized in Supplementary Table S1.

**Figure 1:**
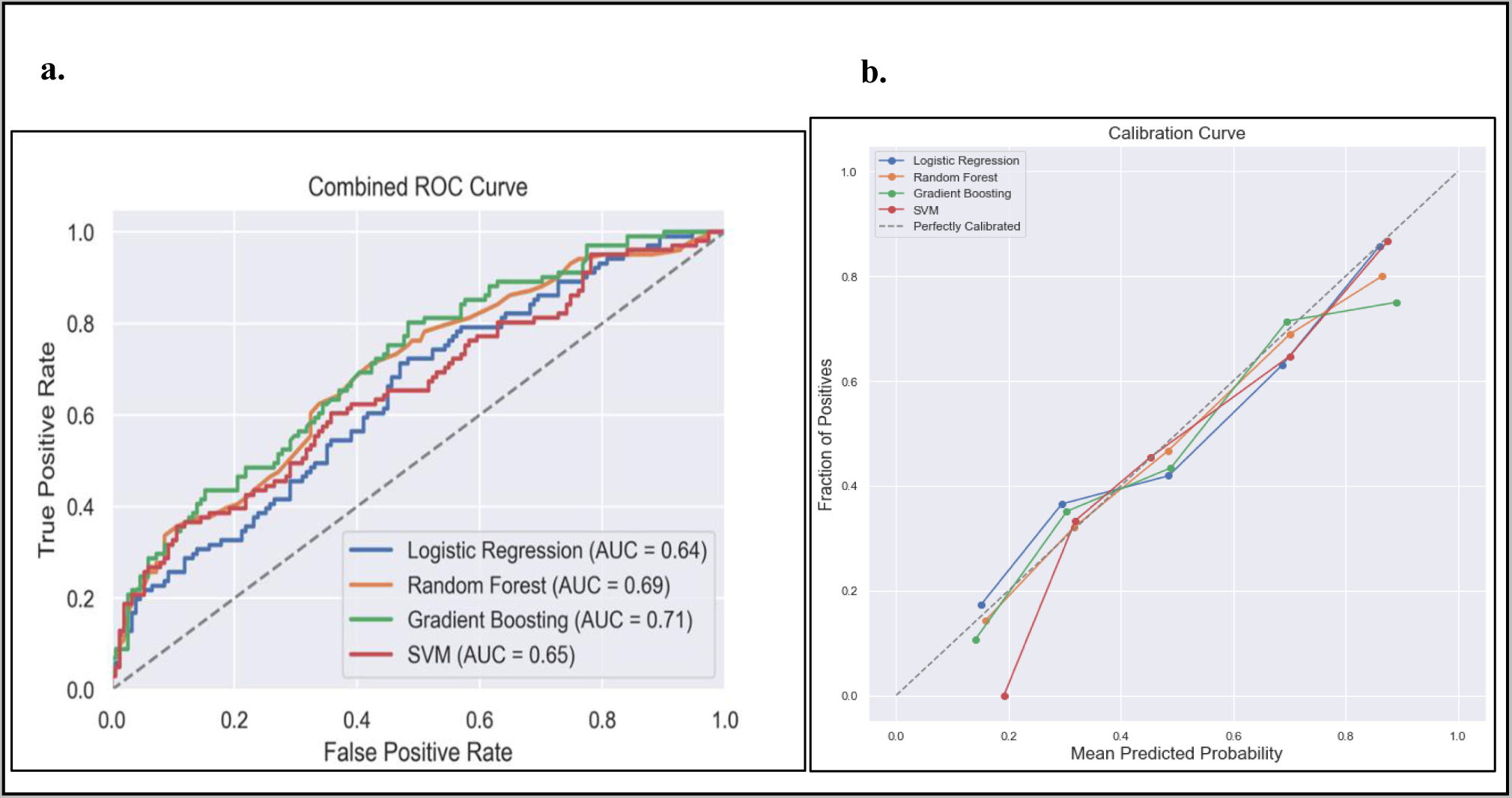
Comparison of the four prediction models (Logistic regression, Random Forest, Gradient Boosting and Support Victor Machine (SVM)) for acute pain intensity prediction. **a.** receiver operating curve, area under the curve (AUC) values for the four models in testing dataset. **b.** calibration curve to compare the mean predicted probability and the fraction of positives for the four models.

**Table 2:** Discrimination metrics and the performance of the models predicting acute pain intensity by the end of RT.

| Model | AUC | Recall | Precision | F1 Score |
| --- | --- | --- | --- | --- |
| Logistic Regression (LR) | 0.6442 | 0.3366 | 0.7881 | 0.4072 |
| Random Forest (RF) | 0.6895 | 0.3762 | 0.8411 | 0.4663 |
| Gradient Boosting (GB) | 0.7085 | 0.3861 | 0.8609 | 0.4845 |
| Support Vector Machine (SVM) | 0.6536 | 0.2277 | 0.9470 | 0.3485 |

SVM classifier demonstrated the highest precision (0.95) but falls behind in sensitivity (0.23) and overall F1 Score (0.35). The calibration evaluated in the testing dataset demonstrated that the RF model exhibits the lowest ECE at 0.0228, suggesting that its predicted probabilities are well-calibrated and closely reflect the true event probabilities. The SVM follows closely with an ECE of 0.0342, indicating good calibration as well. LR also performs well with an ECE of 0.0436, while Gradient Boosting shows a slightly higher ECE at 0.0589. These results suggest that the RF is particularly reliable in providing well-calibrated probability estimates, making them potentially more accurate in capturing the true uncertainty associated with predictions compared to the other models. Calibration plots of the four models are illustrated in (Figure 1.b).

The feature importance analysis from the GBM Classifier was demonstrated in Figure 2, revealed key factors influencing the prediction of the pain intensity by the end of RT. Baseline pre-radiotherapy pain score and changes in weight emerged as the most crucial contributors, emphasizing the significance of the initial pain levels and weight alterations in predicting acute pain by the end of RT (mean importance: 0.244 ± 0.038 and 0.214 ± 0.031, respectively). Additionally, changes in heart rate (i.e., pulse) (mean 0.147 ± 0.028), age (mean 0.123 ± 0.026), and drug abuse (mean 0.055 ± 0.018), exhibit considerable importance. Tumor stage, as represented by clinical T and N, also contribute moderately to the prediction, with mean importance 0.036 ± 0.013 and 0.041 ± 0.016 respectively. The type of primary cancer, treatment approach (i.e., receipt of systemic therapy, surgery), had a moderate impact compared to the sociodemographic factors (race, smoking, alcohol, sex) show lower degrees of importance, (see Figurer 2).

**Figure 2:**
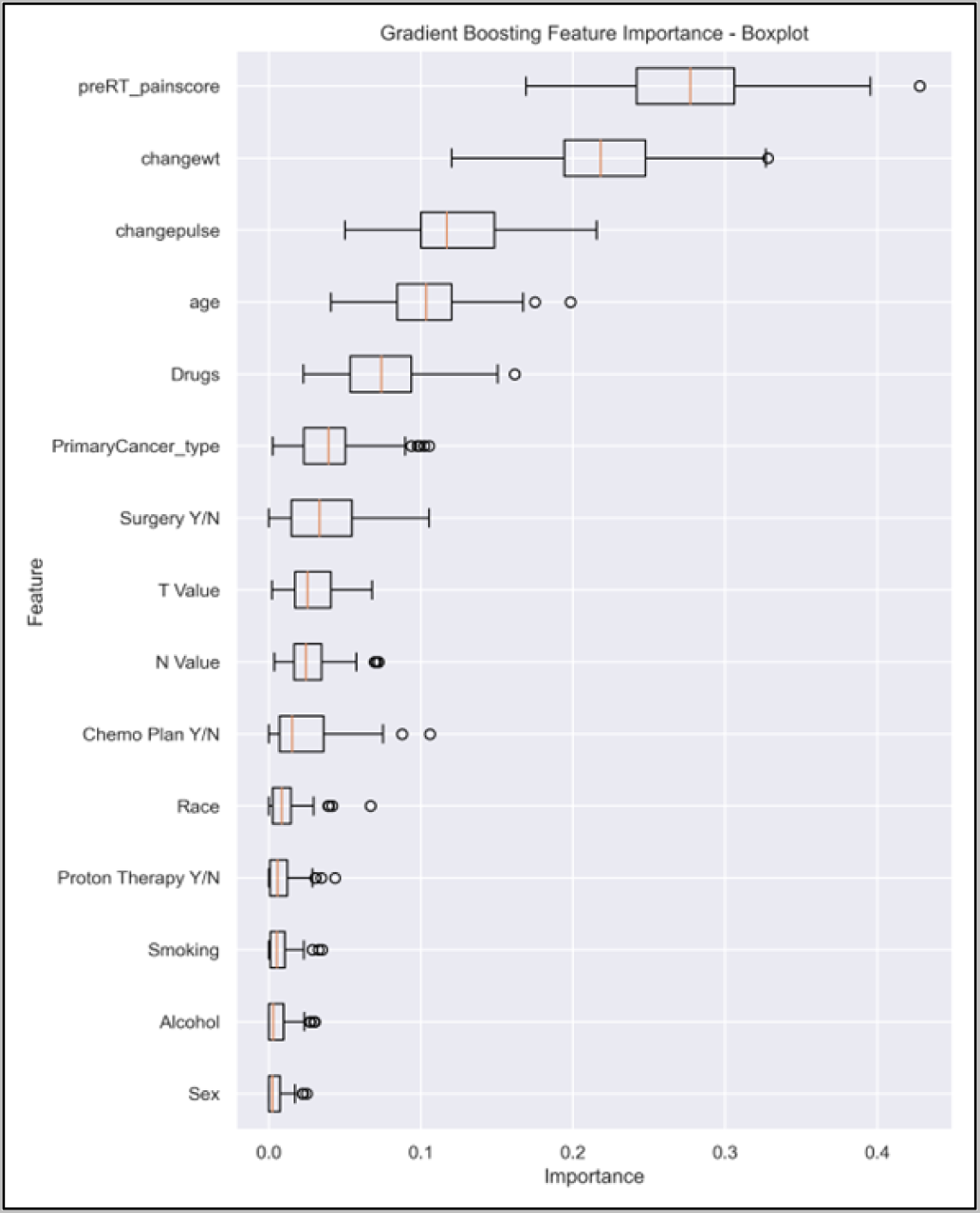
The box plot visually summarizes the distribution of feature importance obtained from a GBM. Pre-RT_painscore: pre-RT pain score; changewt: change in weight; changepulse: change in pulse (HR), Drugs (drug abuse history), primaryCancer_Type: primary cancer type (OC, OPC or unknown primary); Chemo Plan Y/N: chemotherapy plan Yes/No; T value: clinical T stage, N value: clinical N stage, Y/N: Yes or No.

### Models’ development and comparison for predicting total morphine equivalent daily dose (MEDD) at the end of RT in OC/OPC

A total of 838 patients [low MEDD (n= 458, 55%), high MEDD (n=380, 45%)] were included in this analysis after dropping patients with missing data. Results of the discrimination metrics were summarized in Table 3.

**Table 3:** Discrimination metrics and the performance of the models predicting total MEDD by the end of RT.

| Model | AUC | Recall | Precision | F1 Score |
| --- | --- | --- | --- | --- |
| Logistic Regression | 0.6693 | 0.5000 | 0.6196 | 0.5534 |
| Random Forest | 0.6295 | 0.2456 | 0.6512 | 0.3567 |
| Gradient Boosting | 0.5847 | 0.4211 | 0.5217 | 0.4660 |
| Support Vector Machine | 0.6066 | 0.1930 | 0.7333 | 0.3056 |

The performance evaluation results across the four models reveal varying strengths and trade-offs (Table 4). The LR outperformed others in terms of AUC (0.67), indicating its effectiveness in overall predictive performance for predicting the total MEDD at the end of RT (Figure 3.a). RF showed AUC score of (0.63). GBM showed a better balance between precision (0.52) and recall (0.42) while the lowest AUC (0.58). The SVM model achieved the highest precision (0.73) but at the cost of lower recall (0.19). Significant difference in AUC scores was found between LR model and GBM (P=0.007, 95% CI 0.02 to 0.128); AUC-SVM and AUC-GBM (P=0.02, 95% CI −0.126 to −0.011); and AUC-RF and AUC-GBM (P=0.019, 95% CI −0.09 to −0.008), while no significant difference was detected between the AUC scores of other models. DeLong test results were summarized in Supplementary Table S2.

**Figure 3:**
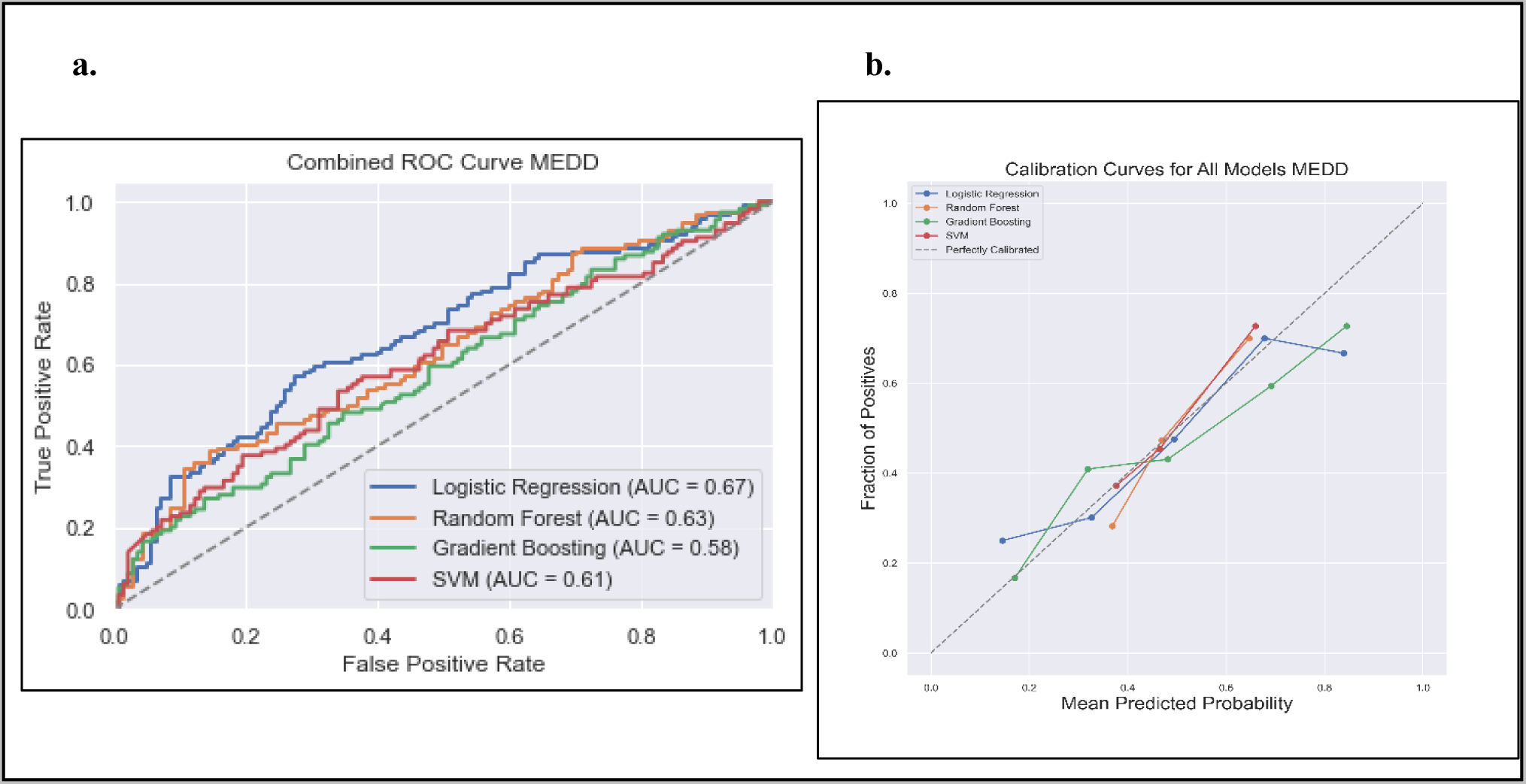
Comparison of the four prediction models (Logistic regression, Random Forest, Gradient Boosting and Support Victor Machine (SVM)) for the total MEDD prediction. **a.** receiver operating curve, area under the curve (AUC) values for the four models in testing dataset. **b.** calibration curve to compare the mean predicted probability and the fraction of positives for the four models.

**Table 4:**
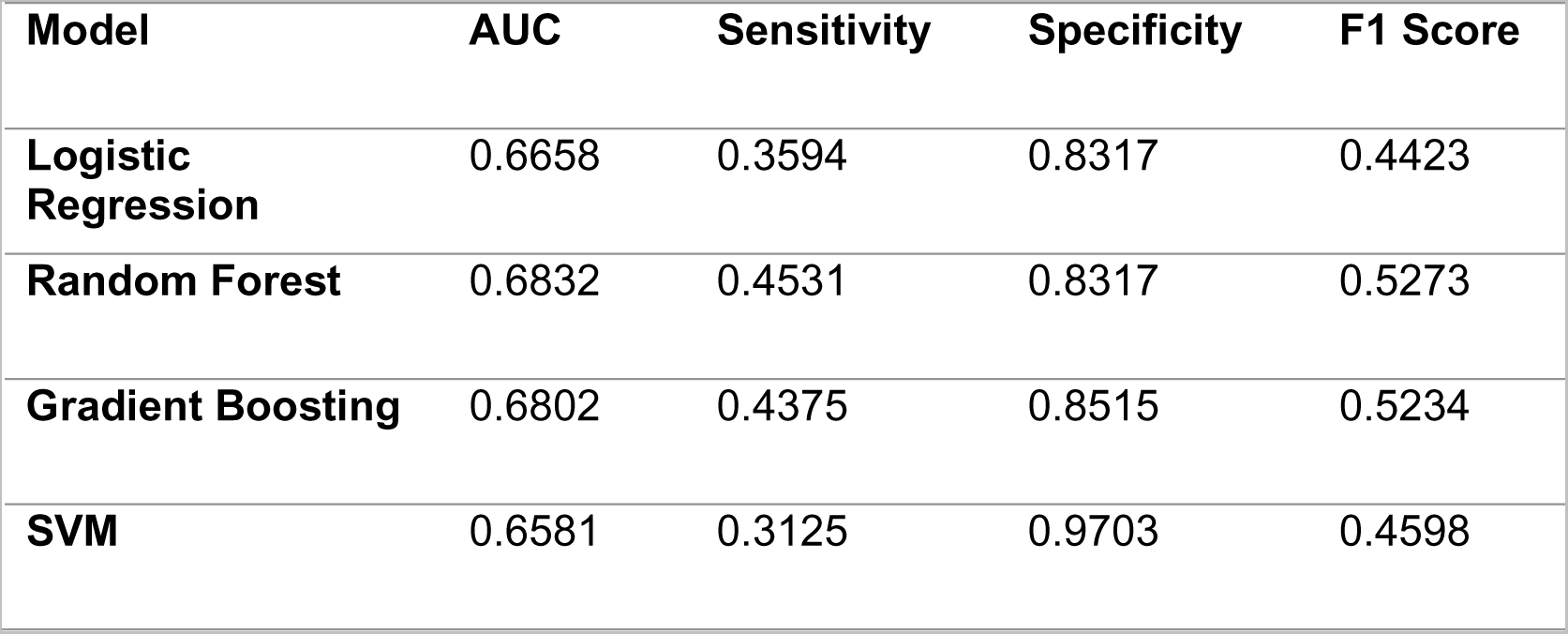
Discrimination metrics and the performance of the models predicting analgesic efficacy by the end of RT.

The calibration analysis of the models revealed notable differences in their ability to provide well-calibrated probability estimates. The RF model emerged as the top-performing model with the lowest Expected Calibration Error (ECE) of 0.0569, indicating a highly accurate alignment between predicted probabilities and actual outcomes. GBM model followed closely with an ECE of 0.0790, revealing good calibration performance. The SVM model exhibits a higher ECE of 0.1588, suggesting a less precise calibration. LR fell in between, with an ECE of 0.0795. These findings highlight the importance of assessing calibration in addition to traditional performance metrics, emphasizing RF as a particularly reliable model in providing well-calibrated probability estimates for the classification task at hand. Calibration plots of the four models were illustrated in (Figure 3.b)

Feature importance results from the logistic regression model permutation analysis, demonstrated in Figure 4, revealed that the most influential features include the “baseline pre-RT pain score” (mean 0.066 ± 0.021), demonstrating a substantial positive association with the target variable. Other positively impactful features, such as clinical T stage (0.01 ± 0.013) and sex (0.009 ± 0.01) contribute positively to the model’s predictions. Conversely, features like drug abuse (−0.009 ± 0.011), alcohol (−0.005 ± 0.01) and age (−0.004 ± 0.014) exhibited negative associations, indicating their potential role in decreasing the likelihood of the target outcome.

**Figure 4:**
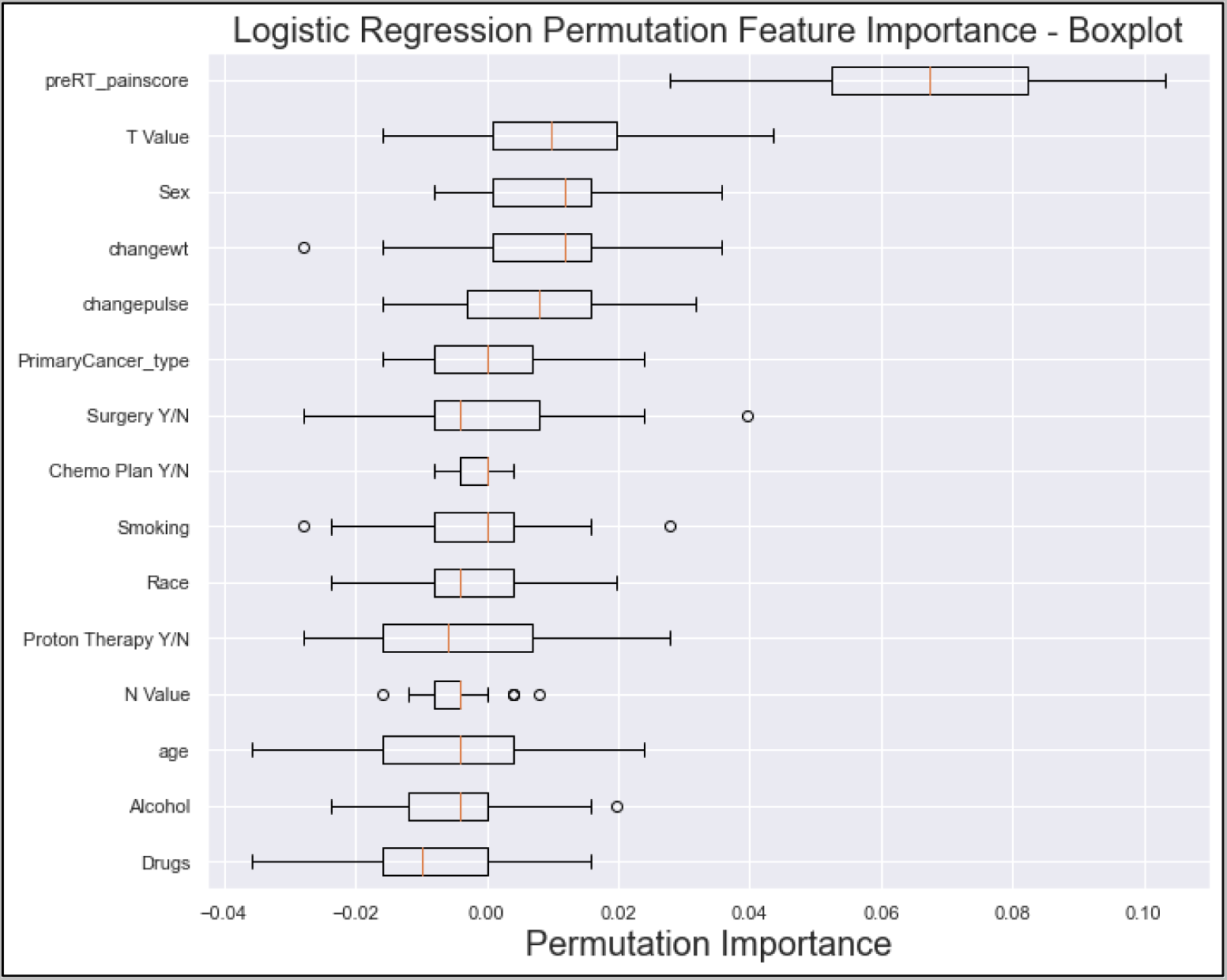
The box plot visually summarizes the distribution of LR permutation feature importance. Pre-RT_painscore: pre-RT pain score; changewt: change in weight; changepulse: change in pulse (HR), Drugs (drug abuse history), primaryCancer_Type: primary cancer type (OC, OPC or unknown primary); Chemo Plan Y/N: chemotherapy plan Yes/No; T value: clinical T stage, N value: clinical N stage, Y/N: Yes or No.

### Models’ development and comparison for predicting analgesic (i.e., opioid) efficacy at the end of RT in OC/OPC

A total of 548 patients [analgesic efficacy (n= 335, 61%), analgesic inefficacy (n=213, 39%)] were included in this analysis after dropping patients with missing data. Results of the discrimination metrics were summarized in Table 4. ROC curves and AUC scores were demonstrated in Figure 5. Analysis of the models’ evaluation results revealed that The LR model achieved an AUC score of 0.67, a sensitivity (recall) of 0.36, a specificity of 0.83, and an F1 score of 0.44. The RF model slightly edged out the other models in AUC score with an AUC of 0.68. It demonstrated better sensitivity (0.45) and maintained a high specificity of 0.83., resulting in an F1 score of 0.53. The GBM performed similarly well with an AUC of 0.68, a sensitivity of 0.44, a specificity of 0.85, and an F1 score of 0.52. The SVM model achieves an AUC of 0.66, a sensitivity of 0.31, a high specificity of 0.97, and an F1 score of 0.46. No significant difference in AUC scores was detected among the four models, (Supplementary Table S3).

**Figure 5:**
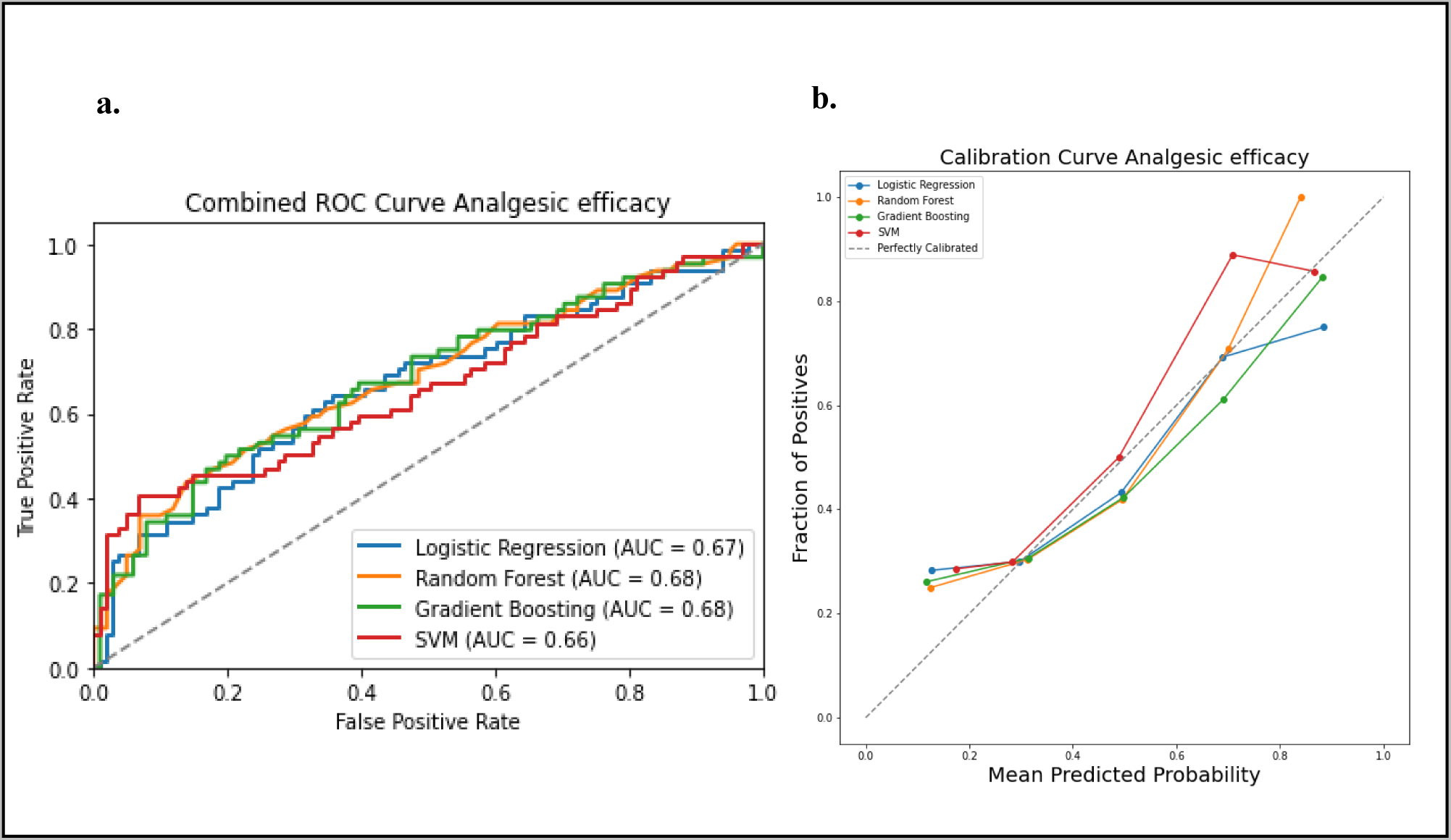
Comparison of the four prediction models (Logistic regression, Random Forest, Gradient Boosting and Support Victor Machine (SVM)) for the analgesic efficacy status. **a.** receiver operating curve, area under the curve (AUC) values for the four models in testing dataset. b. calibration curve to compare the mean predicted probability and the fraction of positives for the four models

Analysis of calibration results revealed that the SVM model stood out with the lowest ECE of 0.0636, suggesting highly accurate and well-calibrated probability predictions. The GBM followed closely with an ECE of 0.0684, indicating good calibration. Meanwhile, the LR and RF models exhibit slightly higher ECE values of 0.0715 and 0.0756, respectively, implying some room for improvement in their calibration performance.

The feature importance results obtained from the RFM, demonstrated in Figure 6, revealed the relative contribution of each feature in predicting the analgesic efficacy. The top features that influenced the model include Baseline pre-RT pain score (0.1696 ± 0.017), change in weight (0.1686 ± 0.01), and change in pulse (0.1565 ± 0.01), indicating their significant impact on the model’s predictions. Other notable features include age (0.1439 ± 0.008), T Value (0.0665 ± 0.005), and N Value (0.0545 ± 0.005). On the other hand, features like the primary cancer type’ (0.0169 ± 0.003), sex (0.0175 ± 0.003), and race (0.0193 ± 0.003) exhibited lower importance.

**Figure 6:**
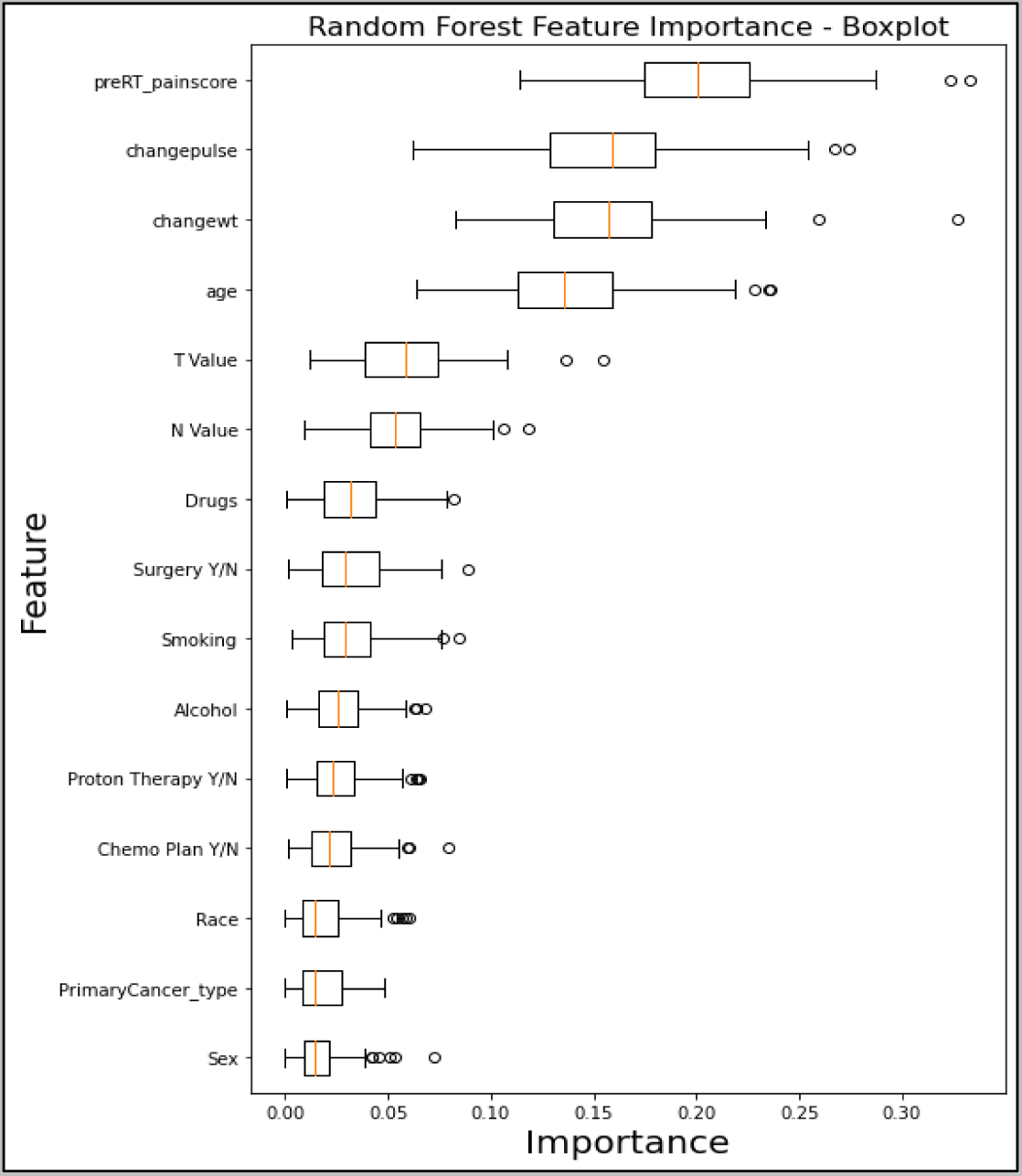
The box plot visually summarizes the distribution of RF feature importance. Pre-RT_painscore: pre-RT pain score; changewt: change in weight; changepulse: change in pulse (HR), Drugs (drug abuse history), primaryCancer_Type: primary cancer type (OC, OPC or unknown primary); Chemo Plan Y/N: chemotherapy plan Yes/No; T value: clinical T stage, N value: clinical N stage, Y/N: Yes or No.

## Discussion

Acute pain is a complex symptom experienced by OC/OPC patients receiving RT. Due to dose-dependent progression of RT-associated toxicities (i.e., oral mucositis, dermatitis, dysphagia), the multifaceted nature of acute pain (i.e., inflammatory, nociceptive, neuropathic components), and lack of data-driven clinical decision support tools to guide pain management, effective pain control is difficult to achieve. This study addresses the existing critical gap in predicting acute pain and opioid usage in OC/OPC patients undergoing RT and identifying the important clinical features affecting pain intensity and opioid usage, using ML. Our results revealed that applying supervised ML predictive models was helpful in early pain prediction and opioid optimization. Among the four models utilized in this study, including GBM, RF, SVM, and LR, GBM emerged as the most promising, exhibiting the highest AUC (0.71), recall (0.39), and F1 score (0.48) for acute pain intensity prediction. LR excelled in predicting total MEDD, displaying competitive AUC scores (0.67). RF demonstrated the highest AUC (0.683) in analgesic efficacy prediction status prediction. These findings emphasize the potential of ML models, particularly GBM and RF, in enhancing risk stratification and prediction accuracy for critical outcomes in OC/OPC patients undergoing RT. Our different results in discrimination metrics highlight the importance of considering specific evaluation metrics based on the clinical requirements.

Model calibration is especially important when deploying ML models for clinical decision-making, as it enhances the reliability of probability. Model calibration estimates a crucial aspect of ML to ensure the predicted probabilities align with the true probabilities of events and poorly calibrated models may provide misleading confidence scores, impacting the interpretability and trustworthiness of predictions [38, 39]. However, few studies focused on evaluation of the calibration of the classification models investigated in the clinical settings [36, 38, 40]. In our study, we generated the calibration curves for the four models to investigate the relationship between the mean predicted probabilities of the positive class and the observed fraction of positive instances. Additionally, we calculated the Expected Calibration Error (ECE) as a key metric used to quantify the calibration performance of a model [36]. Our ECE results revealed good calibration of the four models we investigated for prediction of acute pain intensity, MEDD, and analgesic efficacy in patients with OC/OPC receiving RT.

Although several studies investigated the accuracy and feasibility of ML models for cancer pain prediction and opioids requirement in advance, there is a lack in investigating these models in pain prediction in HNC patients. Chao et al., (2028) explored the role of ML to predict chest wall pain induced by RT in NSCLC patients, using Decision Tress (DT) and RF models build with patient, tumor and dosiomic features. The study demonstrated that ML models are predictive for RT-induced chest wall pain toxicity in lung cancer patients [24]. Additionally, Olling et al., (2018) applied LR, SVM and Generalized Linear Models for prediction pain while swallowing after RT in lung cancer patients and the results demonstrated the effectiveness of ML models in pain prediction during RT in lung cancers [25]. Our study demonstrated the potential effectiveness of ML models in predicting acute pain, opioids dose and analgesic efficacy of OC/OPC patients after RT.

Baseline pain intensity and vital signs were identified as high-risk predictors for cancer-related pain [12, 41]. In a previous study, we established a correlation between vital signs, baseline pain scores, and the intensity of pain during RT for patients with OC/OPC [12]. Uncontrolled pain not only contributes to challenges in chewing and swallowing, leading to weight loss, but also exerts a broader impact on patients’ physiological functions. Elevated pain levels are associated with increased heart rates and changes in blood pressure [12]. Bendall et al., demonstrated an association between vital signs and acute pain [42], additionally Moscato et al., developed an automatic pain assessment tool based on physiological signals recorded by wearable devices [43]. Reyes-Gibby et al., identified the presence of pre-treatment pain as an independent predictor of OC/OPC 5-year survival [44]. According to features importance results, our study highlighted the importance of baseline pre-treatment pain score and the change in vital signs (e.g., weight and heart rate) for contribution in predicting pain intensity, analgesic efficacy, and the total MEDD by GBM and RF models, which is consistent with previous studies [12, 41][43].

The clinical importance of early identification of severe pain in HNCs cannot be overstated. So far, it is extremely hard for clinicians to predict pain severity and identify high risky patients depending on their empirical knowledge. Most clinicians prescribe opioids to OC/OPC patients during therapy according to the pain intensity reported by patients the day of examination, and up to 40% of patients will continue to be dependent on opioids chronically for several months post-therapy [16, 45]. Although following the WHO- analgesic ladder in pain management, pain control during RT in these cancer populations is still challenging and needs further investigations. This study not only demonstrates the predictive capabilities of ML models but also highlights their potential clinical applications. These models can aid in risk stratification, allowing for personalized pain management plans based on individual patient characteristics. By accurately predicting acute pain levels and required opioid doses, clinicians can intervene proactively, mitigating the challenges associated with uncontrolled pain during RT. The integration of AI and ML into clinical practice holds promise for improving outcomes and enhancing the overall well-being and QoL of OC/OPC patients.

### Limitations

Despite the promising findings, this study has limitations that warrant consideration. The retrospective uni-institutional nature of the study and reliance on electronic health records introduce inherent biases. The drop-off of some patients with missing data reduced the size of the final cohort involved in the models, and a bigger multicentric cohort is needed for further validation. The need for external validation of the ML models on independent datasets is crucial to assess their generalizability and robustness. Prospective studies and the incorporation of additional clinical variables may further refine and enhance the predictive performance of these models. We depended our outcome assessment on the patient reported data for pain scoring and the prescription notes in the EHR, however more objective methods are needed for pain assessment and more data about if the patients used the prescribed opioids or not. Recognizing these limitations is imperative for the responsible and effective implementation of AI and ML in clinical settings.

### Conclusion

This research addresses the pressing challenge of predicting acute pain and optimizing opioid usage in patients with OC/OPC undergoing RT. Leveraging ML models, including GBM, RF, SVM, and LR, the study demonstrates the potential effectiveness of these models in enhancing risk stratification and accurately predicting acute pain intensity, total morphine equivalent daily dose (MEDD), and analgesic efficacy at the end of RT. Key predictors identified, such as baseline pain intensity and changes in vital signs, underscore the importance of early identification of high-risk patients, offering the opportunity for proactive and personalized pain management strategies.

## Supporting information

ML_RT pain_Supplementary data

## Data Availability

All data produced are available online at doi: 10.6084/m9.figshare.25114601

## Acknowledgement

We thank The American Legion Auxiliary (ALA) for the ALA Fellowship in Cancer Research, 2022-2024, through The University of Texas, MD Anderson Cancer Center, UTHealth Houston Graduate School of Biomedical Sciences (GSBS). We thank The McWilliams School of Biomedical Informatics at UTHealth Houston, The Student Governance Organization (SGO) Student Excellence award. We thank Daniel GL. Harris from the SMART Core Lab, for helping in the coding process.

## Footnotes

Data sharing statement: An anonymized version of the data set, including clinical variables, pain scores and opioid dose is available at doi: 10.6084/m9.figshare.25114601.

