## Supplementary material for "Comparison of Machine Leaning Models for Prediction of Acute Pain Severity and On-Treatment Opioid Utilization in Oral Cavity and Oropharyngeal Cancer Patients Receiving Radiation Therapy: Exploratory Analysis from a Large-Scale Retrospective Cohort": ML_RT pain_Supplementary data

Supplementary Table S1: DeLong Test results of AUC scores of models predicting acute pain intensity.

| **AUC-Models** | **DeLong p-value** | **95% CI** |
|---|---|---|
| AUC-GBM and AUC-RF | 0.19 | -0.015 to 0.076 |
| AUC-GBM and AUC-SVM | 0.16 | -0.016 to 0.10 |
| AUC-GBM and AUC-LR | 0.09 | 0.696 to 0.644 |
| AUC-RF and AUC-LR | 0.45 | 0.034 to 0.077 |
| AUC-RF and AUC-SVM | 0.67 | -0.042 to 0.07 |
| AUC-SVM and AUC-LR | 0.7 | -0.057 to 0.038 |

Supplementary Table S2: DeLong Test results of AUC scores of models predicting MEDD.

| **AUC-Models** | **DeLong p-value** | **95% CI** |
|---|---|---|
| AUC-GBM and AUC-RF | 0.019 | -0.09 to -0.008 |
| AUC-GBM and AUC-SVM | 0.02 | -0.126 to -0.011 |
| AUC-GBM and AUC-LR | 0.007 | 0.02 to 0.13 |
| AUC-RF and AUC-LR | 0.227 | -0.015 to 0.065 |
| AUC-RF and AUC-SVM | 0.412 | -0.027 to 0.066 |
| AUC-SVM and AUC-LR | 0.809 | -0.037 to 0.048 |

Supplementary Table S3: DeLong Test results of AUC scores of models predicting analgesic efficacy.

| **AUC-Models** | **DeLong p-value** | **95% CI** |
|---|---|---|
| AUC-GBM and AUC-RF | 0.893 | -0.047 to 0.041 |
| AUC-GBM and AUC-SVM | 0.462 | -0.037 to 0.081 |
| AUC-GBM and AUC-LR | 0.532 | -0.031 to 0.059 |
| AUC-RF and AUC-LR | 0.572 | -0.043 to 0.077 |
| AUC-RF and AUC-SVM | 0.405 | -0.034 to 0.084 |
| AUC-SVM and AUC-LR | 0.772 | -0.044 to 0.060 |
